# Cytokines as mediators of the associations of prenatal exposure to phenols, parabens, and phthalates with internalizing behaviours at age 3 in boys: a mixture exposure and mediation approach

**DOI:** 10.1101/2023.03.13.23287180

**Authors:** Olfa Khalfallah, Susana Barbosa, Claire Phillipat, Remy Slama, Cédric Galera, Barbara Heude, Nicolas Glaichenhaus, Laetitia Davidovic

## Abstract

Childhood internalizing disorders refer to inwardly focused negative behaviours such as anxiety, depression, and somatic complains. Interactions between psychosocial, genetic, and environmental risk factors adversely impact neurodevelopment and can contribute to internalizing disorders. While prenatal exposure to single endocrine disruptors (ED) is associated with internalizing behaviours in infants, the associations with prenatal exposure to ED in mixture remain poorly addressed. In addition, the biological mediators of ED mixture effects on internalizing behaviours remain unexplored. ED do not only interfere with endocrine function, but also with immune function and inflammatory processes. Based on this body of evidence, we hypothetised that inflammation at birth is a plausible biological pathway through which ED prenatal exposure could operate to influence offspring internalizing behaviours. Based on the EDEN birth cohort, we investigated whether ED mixture exposure increased the odds of internalizing disorders in 459 boy infants at age 3, and whether the pro-inflammatory cytokines IL-1β, IL-6, and TNF-α measured at birth are mediators of this effect. To determine both the joint and individual associations of ED prenatal exposure with infant internalizing behaviours and the possible mediating role of cytokines, we used the counterfactual hierarchical Bayesian Kernel Machine Regression (BKMR) regression-causal mediation analysis. We show that prenatal exposure to a complex ED mixture has limited effects on internalizing behaviours in boys at age 3. We also show that IL-1β, IL-6, and TNF-α are unlikely mediators or suppressors of ED mixture effects on internalizing behaviours. Further studies on larger cohorts are warranted to refine the deleterious effects of ED mixtures on internalizing behaviours and identify possible mediating pathways.

## BACKGROUND

Alterations in early neurodevelopmental processes can enhance one child’s vulnerability to psychological and emotional distress, and lead to inwardly focused negative behaviours such as internalizing disorders (Liu et al., 2011). Internalizing disorders altogether affect up to 20% of pre-schoolers and are associated with childhood depression and anxiety disorders, peer relationship problems, and somatic symptoms (Liu et al., 2011). Psychosocial, genetic, and environmental factors are potential contributors to internalizing disorders (Tien et al., 2020). Notably, antenatal exposure to environmental pollutants (present in air, food, water, personal care, and household goods) can impair neurodevelopment and lead to internalizing behaviours (Grandjean and Landrigan, 2014). Some of these pollutants are known as endocrine disruptors (ED) since they interfere with hormonal systems. ED are also suspected to exert neurodevelopmental toxicity, with consequences on offspring behavioural outcomes and psychopathology (Grandjean and Landrigan, 2014).

Numerous epidemiological studies have focussed on single ED prenatal exposures and identified candidate ED, mostly phenols, parabens, and phthalates, with consistent associations with internalizing behaviours (Roen et al., 2015) or associated symptoms (emotional and peer relationship problems, somatic complains) (Daniel et al., 2020; Engel et al., 2010; Philippat et al., 2017). Notably, mono-n-butyl-phtalate (mnBP) and monobenzyl phthalate (mBzP) were positively associated with internalizing behaviour at age 3 in boys (Philippat et al., 2017).

ED are rarely present as single compounds, but rather co-occur in mixture. An emerging literature is now addressing the joint effects of ED prenatal exposure on neurodevelopmental disorders (e.g. developmental delay, language delay, or autism spectrum disorders (ASD)) or psychopathology associated with internalizing behaviours). Associations between exposure to a phenol/paraben/phthalate mixtures with developmental delay were reported (Barkoski et al., 2019; Bennett et al., 2022). Also, exposure to organophosphorus pesticides in mixture increased the odds of autism spectrum disorders (ASD) (Bennett et al., 2022). Another study showed that exposure to an ED mixture comprising bisphenol A (BPA), triclosan (Tc), and phthalates, increased the odds of language delay in 30-month-old infants (Caporale et al., 2022). Exposure to a mixture of phthalates, bisphenols, and organophosphorus pesticides was associated with lower nonverbal intellectual quotient at age 6, but not with internalizing behaviour in 782 children from the Generation R cohort (van den Dries et al., 2021). Finally, one study found associations between *in utero* exposure to a mixture of phenols and phthalates and child’s internalizing behaviours, but mainly in girls (Guilbert et al., 2021). These studies suggest a possible effect of ED mixture prenatal exposure on internalizing behaviour and related neurodevelopmental disorders in infants.

To reinforce causal inferences from epidemiological association studies and identify targets for prevention, mediation studies could help refine the biological pathways through which ED mixtures operate. *In vitro* and animal studies have shown that ED can directly interfere with the neuroendocrine system, in particular thyroid or sexual hormones signalling or metabolism, causing disruption of neuronal differentiation and maturation (Lucaccioni et al., 2021). Also, mounting evidence from *in vitro* and animal studies support the notion that ED can also impact immune function. Notably, bisphenols/phenols, phthalates, Tc, and parabens impact the development and functions of most immune cell types (including monocytes, neutrophils, lymphocytes, dendritic cells), with direct effects on the balance in proinflammatory and anti-inflammatory cytokines secreted by immune cells (Nowak et al., 2019). ED are thought to either suppress or enhance immune responses and ED exposure is associated with inflammation, allergies, or autoimmune disorders (Bansal et al., 2018). Cytokines produced by immune cells are key modulators of inflammatory processes and in particular the pro-inflammatory cytokines interleukin (IL)-1β, IL-6, and Tumor Necrosis Factor (TNF)-α. Besides their role in inflammation, these cytokines also regulate essential neurodevelopmental processes, including neurogenesis, synaptogenesis, and myelination (Zengeler and Lukens, 2021). This supports the notion that cytokines can influence the risk of psychopathology, including internalizing behaviours, later in life.

Based on the above, we hypothesised that inflammation at birth, using the cytokines IL-1β, IL-6, and TNF-α measured in cord blood as proxies, is a plausible biological pathway through which ED prenatal exposure could operate to influence offspring’s neurodevelopment, and in particular internalizing behaviours. Relying on the EDEN mother-child cohort, we used Bayesian Kernel Machine Regression-Causal Mediation Analysis (BKMR-CMA) to investigate at once whether antenatal exposure to a mixture of 20 ED increased the odds of internalizing behaviours in boy infants at age 3 and whether IL-1β, IL-6, and TNF-α at birth are mediators of this effect.

## 2. MATERIAL & METHODS

### 2.1. Study sample

The present study is nested within the EDEN mother-child cohort (Heude et al., 2016). Pregnant women were recruited before 24 weeks of gestation in the French University Hospitals of Nancy and Poitiers. Clinical and psychosocial data were gathered from medical records, interviews with the mother and auto-questionnaires. Our study sample consisted of 459 mother-boy pairs and 400 pairs for which cord blood sera, maternal urinary levels of ED during pregnancy, and the behavioural outcome at age 3 were available (Tab. 1).

**Tab. 1.** Study sample characteristics. BMI, body mass index; SD, standard deviation; Min., minimum; Max., maximum; N/A, non-available (N/A) values; Min., minimum; Max., maximum.

|  | Nb (%) | NA (%) | Min. | Q25 | Mean | Median | Q75 | Max. | SD |
| --- | --- | --- | --- | --- | --- | --- | --- | --- | --- |
| Center : Nancy | 212 (46) | 0 |  |  |  |  |  |  |  |
| Center : Poitiers | 247 (54) |  |  |  |  |  |  |  |  |
| Pre-pregnancy BMI (kg/m<sup>2</sup>) | 454 (99) | 1.1 | 15.8 | 20 | 23 | 22 | 25.2 | 44.6 | 4.4 |
| Maternal dietary pattern during pregnancy : Healthy | 390 (85) | 15 | -1.8 | -0.6 | 0.02 | -0.06 | 0.5 | 5.5 | 0.9 |
| Maternal dietary pattern during pregnancy : Western | 390 (85) | 15 | -3.6 | -0.7 | -0.06 | -0.1 | 0.5 | 4.1 | 1 |
| Smoking (nb cigarettes/day) | 450 (98) | 2 | 0 | 0 | 1.1 | 0 | 0 | 16.7 | 2.7 |
| Caffeine consumption during pregnancy (mg/day) | 448 (98) | 2.4 | 0 | 14.2 | 94.4 | 60.8 | 118.5 | 537.3 | 107.3 |
| Alcohol drinking during pregnancy (mean nb glasses/week) | 390 (85) | 15 | 0 | 0 | 0.49 | 0 | 0.3 | 14 | 1.2 |
| Maternal depression during pregnancy (yes) | 102 (22) | 0.7 |  |  |  |  |  |  |  |
| Maternal depression during pregnancy (no) | 354 (77) |  |  |  |  |  |  |  |  |
| Maternal prenatal anxiety (STAI scale) | 458 (99) | 0.2 | 0 | 3 | 9.972 | 7 | 14 | 53 | 9.7 |
| Age at delivery (years) | 459<br>(100) | 0 | 17 | 26 | 29.7 | 29 | 33 | 44 | 4.9 |
| Delivery mode : Vaginal (Reference) | 348 (76) |  |  |  |  |  |  |  |  |
| Delivery mode : Forceps | 50 (11) | 0 |  |  |  |  |  |  |  |
| Delivery mode : C-section | 61 (13) |  |  |  |  |  |  |  |  |
| Trimester of birth : 1<sup>st</sup> | 114 (25) |  |  |  |  |  |  |  |  |
| Trimester of birth : 2<sup>nd</sup> | 129 (28) | 0 |  |  |  |  |  |  |  |
| Trimester of birth : 3<sup>rd</sup> | 120 (26) |  |  |  |  |  |  |  |  |
| Trimester of birth : 4<sup>th</sup> | 96 (21) |  |  |  |  |  |  |  |  |
| Sex: Male | 459<br>(100) | 0 |  |  |  |  |  |  |  |
| Sex: Female | 0 |  |  |  |  |  |  |  |  |
| Parental Separation : yes | 38 (8) | 9.6 |  |  |  |  |  |  |  |
| Parental Separation : no | 377 (82) |  |  |  |  |  |  |  |  |
| Maternal education (years) | 456 (99) | 0.7 | 9 | 12 | 13.9 | 14 | 17 | 17 | 2.6 |
| Paternal education (years) | 416 (91) | 9.4 | 9 | 11 | 13.2 | 12 | 17 | 17 | 2.6 |
| House Income at age 3 | 395 (86) | 13.9 | 2 | 4 | 5.3 | 5 | 6 | 8 | 1.5 |
| Number of siblings | 397 (86) | 13.5 | 0 | 0 | 0.9 | 1 | 1 | 5 | 0.9 |
| Breastfeeding : No | 110 (24) |  |  |  |  |  |  |  |  |
| Breastfeeding : ≤ 6 months | 255 (56) | 0 |  |  |  |  |  |  |  |
| Breastfeeding : > 6 months | 94 (20) |  |  |  |  |  |  |  |  |
| Child's BMI at age 3 (kg/m<sup>2</sup>) | 396 (86) | 13.7 | 12.9 | 15.1 | 15.8 | 15.8 | 16.4 | 21.5 | 1.1 |
| Child's dietary pattern at age 3 : Processed and fast foods | 400 (87) | 12.9 | -1.8 | -0.6 | 0.08 | -0.1 | 0.5 | 12 | 1.2 |
| Child's dietary pattern at age 3 : Recommended guidelines | 400 (87) | 12.9 | -4.1 | -0.7 | -0.08 | -0.02 | 0.5 | 3.6 | 1.1 |
| Age at internalizing behaviour score assessment (years) | 403 (88) | 12.2 | 1061 | 1135 | 1153.3 | 1148 | 1163.5 | 1411 | 30.5 |
| Internalizing behaviour score | 400 (87) | 12.9 | 0 | 1 | 3 | 3 | 4 | 15 | 2.4 |

### 2.2. Outcome: child’s internalizing behaviour at age 3

Child’s behaviour was assessed by the Strengths and Difficulties Questionnaire (SDQ) completed by the mother (Goodman, 1997). The SDQ is a widely used psychometric instrument to evaluate psychopathology in children (Goodman, 2001). This questionnaire includes 25 items scored on a 3-point scale (0: not true; 1: somewhat true; 2: certainly true) that are combined into 4 difficulties sub-scores: conduct problems, hyperactivity-inattention, peer relationship problems and emotional symptoms, and 1 strength sub-score: prosocial behaviour. Each sub-score ranges from 0 to 10. The internalizing behaviour scale, refers to problems of withdrawal, somatic complaints, and anxiety/depression. The internalizing behaviour score is computed by summing the peer relationship problems and emotional symptoms sub-scores (Goodman, 2001).

### 2.3. Exposure

Women were asked to collect first morning urine void at home just before the prenatal study visit between 22 and 29 gestational weeks. Using solid phase extraction-high-performance liquid chromatography-isotope dilution tandem mass spectrometry, we measured 20 ED in maternal urine (Tab. 2): five phenols (2,4-dichlorophenol (2,4-DC); 2,5-dichlorophenol (2,5-DC); BPA; benzophenone-3 (Bzp-3); Tc), four parabens (butyl-paraben (BP); ethyl-paraben (EP); propyl-paraben (PP); methyl-paraben (MP)), seven phthalates (monoethyl phthalate (mEP); mono-n-butyl phthalate (mnBP); mono-isobutyl phthalate (miBP); monobenzyl phthalate (MBzP); monocarboxynonyl phthalate (mCNP); monocarboxyoctyl phthalate (mCOP), mono(3-carboxypropyl) phthalate (mCPP)), and four di(2-ethylhexyl) phthalate (DEHP) metabolites (mono(2-ethylhexyl) phthalate (mEHP); mono(2-ethyl-5-hydroxy-hexyl) phthalate (mEHHP); mono(2-ethyl-5-oxohexyl) phthalate (mEOHP); mono(2-ethyl-5-carboxypentyl) phthalate (mECPP)).

**Tab. 2.**
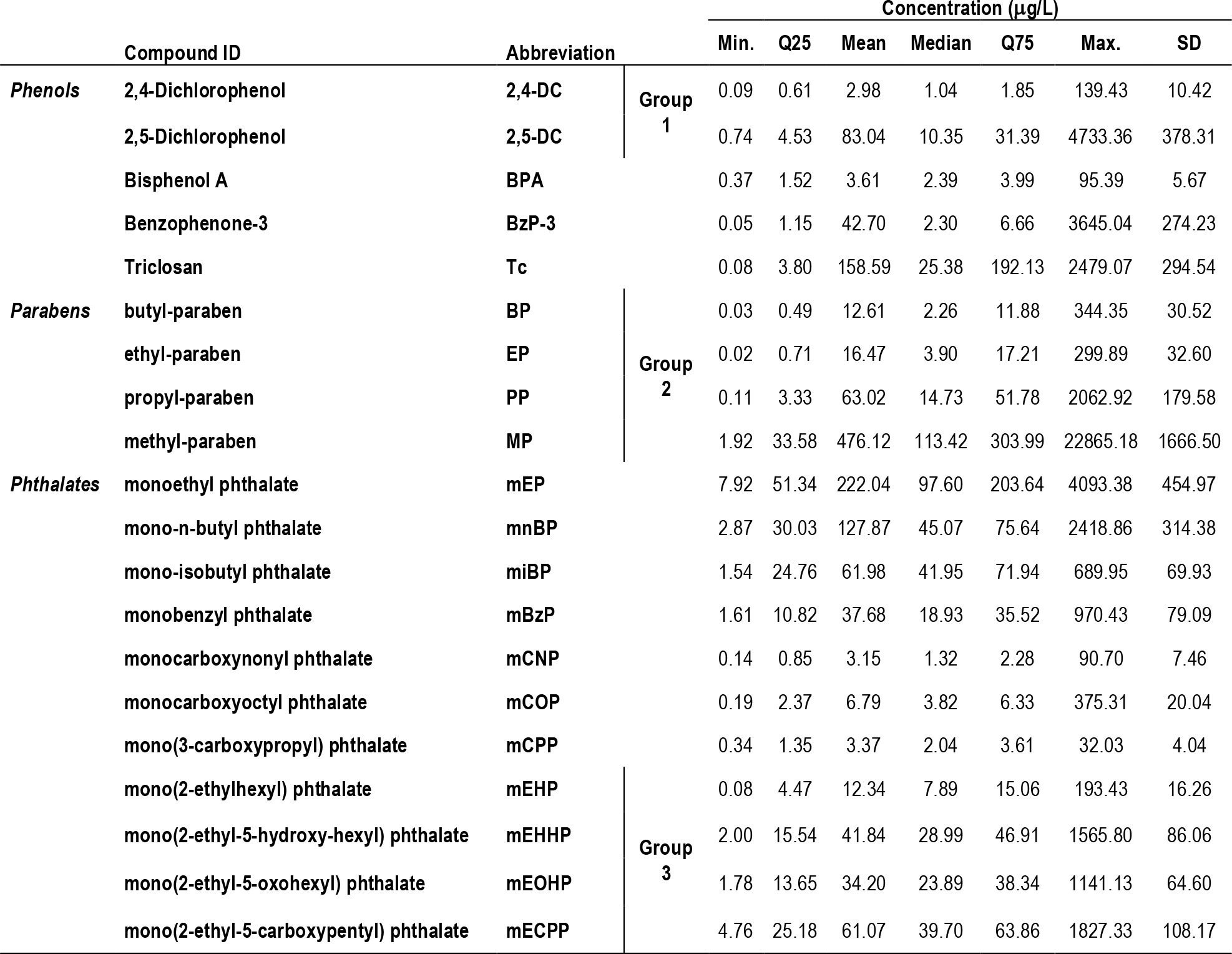
ED concentrations in maternal urine. Min., minimum; Max., maximum; Min., minimum; Max., maximum; SD, standard deviation.

Urinary creatinine was used as an internal standard for normalization. Measures were performed at the Centers for Disease Control and Prevention laboratory (Atlanta, Georgia, USA), according to established procedures (Silva et al., 2007; Ye et al., 2005). We used instrumental reading values for biomarker concentrations below the limits of detection. Instrumental reading values equal to 0 (i.e., indicative of no signal) were replaced by the lowest instrumental reading value divided by the square root of 2. Prior to statistical analysis, concentrations were standardized for creatinine, sampling conditions (e.g., hour of sampling, gestational age at sampling), and analysis year (2008 or 2011) using a two-step standardization approach (Mortamais et al., 2012). Standardized concentrations were then log 2-transformed for downstream analyses.

### 2.4. Cytokine measures in cord blood samples

Venous cord blood was sampled immediately after birth and allowed to clot. Blood samples were centrifuged within 24 h post collection. Serum was collected, aliquoted, and stored at −80°C. Cord blood sera were assessed for the levels of 28 cytokines (Barbosa et al., 2020), using a multiplex electrochemiluminescence-based immunoassay and according to the manufacturer’s recommendations (Meso Scale Diagnostics, Rockville, USA). Details related to procedures for measurement, quality control, and coefficient of variations are provided in (Barbosa et al., 2020). As proxies for inflammation at birth, we considered the emblematic pro-inflammatory cytokines IL-1β, IL-6, and TNF-α (Tab. 3). For cytokine concentrations below the lower limit of detection (LLOD), half the LLOD value was imputed as recommended for immunological measurements constrained by detection limits (Uh et al., 2008).

**Tab. 3.** Pro-inflammatory cytokines concentrations in cord blood serum. Min., minimum; Max., maximum; Min., minimum; Max., maximum; SD, standard deviation.

| Cytokine | Min. | Mean | Median | Max. | SD |
| --- | --- | --- | --- | --- | --- |
| IL-1β (pg/ml) | 0.02 | 1.66 | 0.20 | 456.20 | 21.33 |
| IL-6 (pg/ml) | 0.16 | 23.89 | 2.77 | 2756.37 | 167.15 |
| TNF-α (pg/ml) | 1.44 | 4.77 | 3.20 | 631.53 | 29.34 |

### 2.5. Covariates

Previous studies have identified maternal, perinatal and psychosocial variables associated with behavioural abnormalities in children (de La Rochebrochard and Joshi, 2013; Gumusoglu and Stevens, 2019; Melchior et al., 2015; Noonan et al., 2018; Polanska et al., 2015; van der Waerden et al., 2015). Based on these studies, and considering that many of these variables can also influence cytokine production or ED exposure, we selected the following variables (Fig. 1). Maternal variables were: maternal age at delivery (years), maternal pre-pregnancy body mass index (BMI in kg/m^2^), alcohol drinking during pregnancy (mean number of glasses/week), smoking during pregnancy (number of cigarettes/day), caffeine intake during pregnancy (mg caffeine/day), maternal dietary pattern during pregnancy (Healthy, vs. Western), record of depression during pregnancy (yes/no), and symptoms of prenatal anxiety. Maternal dietary patterns were constructed based on frequency food diaries validated in previous studies led in the EDEN cohort (Collet et al., 2021; Galera et al., 2018; Yuan et al., 2017). Maternal depression was assessed at 24-28 weeks of amenorrhea, using the Center for Epidemiological Studies Depression questionnaire (CES-D (Radloff, 1991)) and women presenting a CES-D score above a cutoff of 17 were considered as depressed. Symptoms of maternal anxiety were assessed at 24-28 weeks of amenorrhea, using the State-Trait Anxiety Inventory (STAI) (Spielberger and Vagg, 1984) and the continuous STAI score was used to assess the severity of anxiety symptoms. Perinatal variables were: delivery mode (vaginal, forceps, C-section), birth trimester, and recruitment center (Nancy or Potiers). Gestational age or birth weight were not included as covariates as they could be on the path from ED exposure to internalizing behaviours and contribute to lower estimated effects. Postnatal socioeconomic variables were: house income at age 3 (in €), maternal and paternal education duration (years), parental separation (yes/no), and breastfeeding (no, < 6 months, > 6 months). Child’s variables at age 3 were: BMI (kg/m^2^) and child’s diet (Processed and fast foods vs. Recommended food guidelines, according to a food frequency diary validated in previous studies led in EDEN cohort (Deschamps et al., 2009; Lioret et al., 2015; Yuan et al., 2017)), Multiparity (number of older siblings) and age at SDQ assessment (years) were also included.

**Fig. 1.**
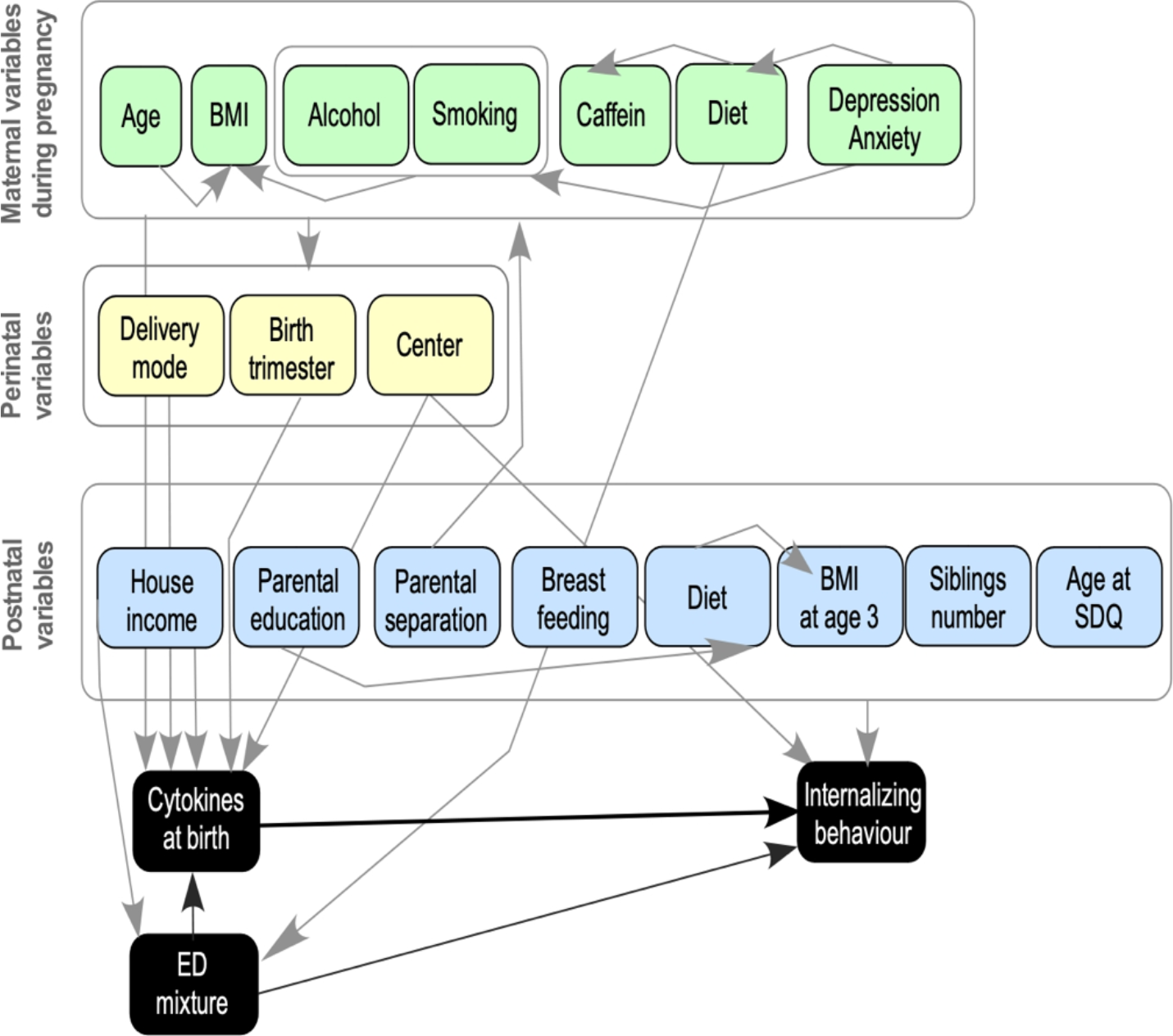
Relationships between covariates, confounders, exposure (ED mixture), mediators (cytokines at birth), and outcome (internalizing behaviour score).

### 2.6. Statistical analyses

#### Multiple imputation (MI) of missing data

The following analysis were performed over 30 multiple-imputed datasets generated using the mice package in R (van Buuren and Groothuis-Oudshoorn, 2011).

#### Bayesian Kernel Machine Regression causal mediation analysis (BKMR-CMA) framework

Until recently, epidemiological studies to study ED effects in complex mixtures were limited by available statistical methodologies to handle i) multiple and correlated ED exposures, ii) non-linear effects of ED exposures on the outcome, iii) regression instability as the number of exposures and covariates increases (Demeinex and Slama, 2019; Tanner et al., 2020). Here we relied on the BKMR-CMA framework which now allows to simultaneously estimate the total effect of ED mixture exposure and mediating effects (Devick et al., 2022). BKMR appears currently as the least biased and most accurate in estimating independent and cumulative effects of chemicals, as well as dose-response relationships and interaction effects (Bobb et al., 2018). The BKMR-CMA approach defines natural direct effects (NDE) and natural indirect effects (NIE) that sum up to the total effect (TE) (Fig. 2).

**Fig. 2.**
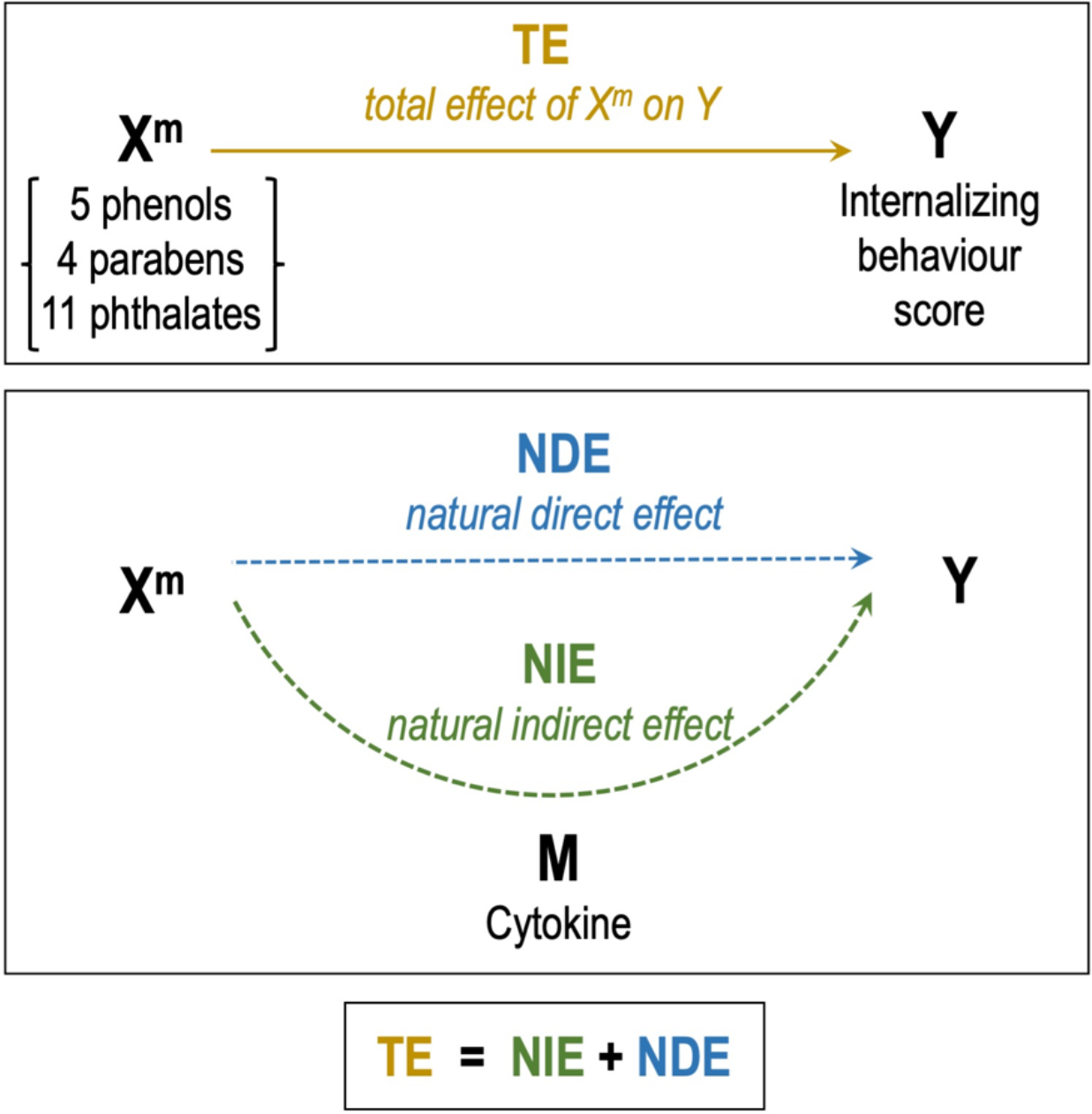
BKMR causal mediation analysis counterfactual framework. Relationships between mixture exposure (X^m^), outcome (Y), and mediator (M).

In a counterfactual framework, the individual causal effect of the exposure on the outcome is defined as the hypothetical contrast between the outcomes that would be observed in the same individual at the same time with or without the exposure, or in the presence of two different levels of the exposure. The NIE measures the average difference in counterfactual outcomes when fixing the exposure to level z, while the mediator varies from the level it would have taken if the exposure was set to z compared to z*. The NDE captures the average difference in the counterfactual outcomes for a change in exposure level z* to z, while fixing the mediator to the level it would have taken if the exposure was set to z*.

Because the BKMR-CMA framework does not allow for considering multiple mediators in the same model, we conducted the mediation analysis using each cytokine independently. We simulated counterfactuals to estimate TE, NDE and NIE for a change in the exposure from z* to z, where each ED is set at the 25^th^ and 75^th^ percentile, while each cytokine is set at the 25^th^, 50^th^ or 75^th^ percentile. BKMR-CMA analysis was performed according to (Devick et al., 2022), using the R statistical analysis software using the ‘bkmr’ package version 0.2.0 (Bobb, 2022). All numerical variables (exposure, outcome and covariates) were log2-transformed and scaled. To account for high within-group correlation but relatively lower across-group correlations across ED, we used the hierarchical variable selection option in the bkmr package. Group 1 consisted of 2,4-DC and 2,5-DC, group 2 of the 4 parabens (MP, EP, BP, PP), Group 3 of the four DEHP metabolites (mEHP, mEHHP, mEOHP, mECPP), while the remaining 10 ED were considered individually (Tab. 2). For each MI dataset, we conducted 30,000 iterated BKMR runs and produced estimates for each MI dataset with 95% credible intervals (CI). In Baeysian statistics, the 95% CI is analogous to the confidence interval in non-Baeysian statistics. The 95% CI determines the distribution of possible estimate values between its upper and lower boundaries with the subjective probability of 95%. We derived from BKMR-CMA runs the Posterior Inclusion Probabilities (PIP), the full posterior distribution of TE, NDE, and NIE estimates and 95% CI, the predictor-response functions, and the contribution of individual predictors to the response.

### 2.7. Ethics

The EDEN cohort received approval from the Ethical Research Committee (CCPPRB) of Bicêtre Hospital and from the French National Data Protection Agency (CNIL). Informed written consent was obtained from parents at the time of enrolment and after delivery. The analysis of blinded urine specimens at CDC laboratories was determined not to constitute engagement in human subjects’ research.

## 3. RESULTS

Our study sample consisted of 459 mother-boy pairs recruited in two maternity centres (Tab.1). As for exposure, we considered 20 ED metabolites in mixture. Among these 20 ED, ten (BPA, Tc, mEP, mnBP, mBzP, mEHP, mEHHP, mEOHP, mECPP, mCOP) were present in an ED mixture recently shown to be associated with language delay in young infants (Caporale et al., 2022). Our mixture also contains parabens known to co-occur with phenols in ED mixtures (Bennett et al., 2022). All 20 ED were measured and systematically detected in maternal urinary samples collected during pregnancy among mothers who delivered a boy (Philippat et al., 2017) (Table 2). Tc, MP, mEP, and mnBP presented the highest mean concentrations (> 100 μg/L), while 2,4-DC, BPA, mCNP, and mCPP presented the lowest (<5 μg/L) (Tab. 2). Medium to strong correlations were observed among ED from the same classes (Fig. S1), in particular among the two dichlorophenols (2,4-DC, 2,5-DC), the four parabens (MP, EP, PP, BP), and four DEHP metabolites (mEHP, mEHHP, mEOHP, mECPP). Weak correlations were observed across ED classes, with Tc correlating with 2,4-DC, parabens and one phthalate metabolite mEP, and BPA correlating with phthalates and DEHP metabolites (Fig. S1). As mediators, we considered the cytokines IL-1β, IL-6, and TNF-α which were measured in cord blood serum samples collected at birth (Barbosa et al., 2020) (Tab. 3). IL-1β, IL-6, and TNF-α levels correlated in cord blood serum samples (Fig. S2) and these cytokines are induced in inflammatory conditions.

We used hierarchical BKMR-CMA to model at once the effects of the 20 ED mixture on internalizing behaviour and the possible mediating effects of cytokines. Hierarchical BKMR allowed to group ED metabolites from the same class showing strong correlations (Group1: 2,4-DC, 2,5-DC; Group 2: BP, EP, PP, MP; Group 3: mEHP, mEHHP, mEOHP, mECPP), while uncorrelated ED metabolites were considered independently. When addressing the total effect (TE) of ED mixture across BKMR runs, a joint change in the ED mixture from the 25^th^ percentile value to its 75^th^ percentile value was associated with an increase of 1.018 on the internalizing behaviour score (Tab. S1). However, the wide CI comprising zero [-1.822; 3.858] (Tab. S1) was indicative of an instability in the estimation.

The PIP provided further insights on the ED selection across BKMR runs and therefore of the importance of each ED metabolite or metabolite group in defining the exposure-outcome association (Fig. 3). In general, the estimated individual PIP were small (PIP<0.5), with the phthalate miBP exhibiting the highest value (PIP>0.2). PIP inspection also highlighted possible minor contribution of mEP and mnBP in defining the association between ED exposure and internalizing behaviour scores at age 3. The importance of miBP and mnBP was also confirmed by examining individual dose-response associations for each ED (Fig. 4), when all the other ED were fixed at their median. miBP somehow presented with a non-linear negative relationship with the outcome, while mnBP exhibited a positive linear relationship with the outcome. The associations of the other ED with internalizing behaviour scores were mainly unchanged while holding the other compounds at their median, indicating no synergistic or multiplicative interactions.

**Fig. 3.**
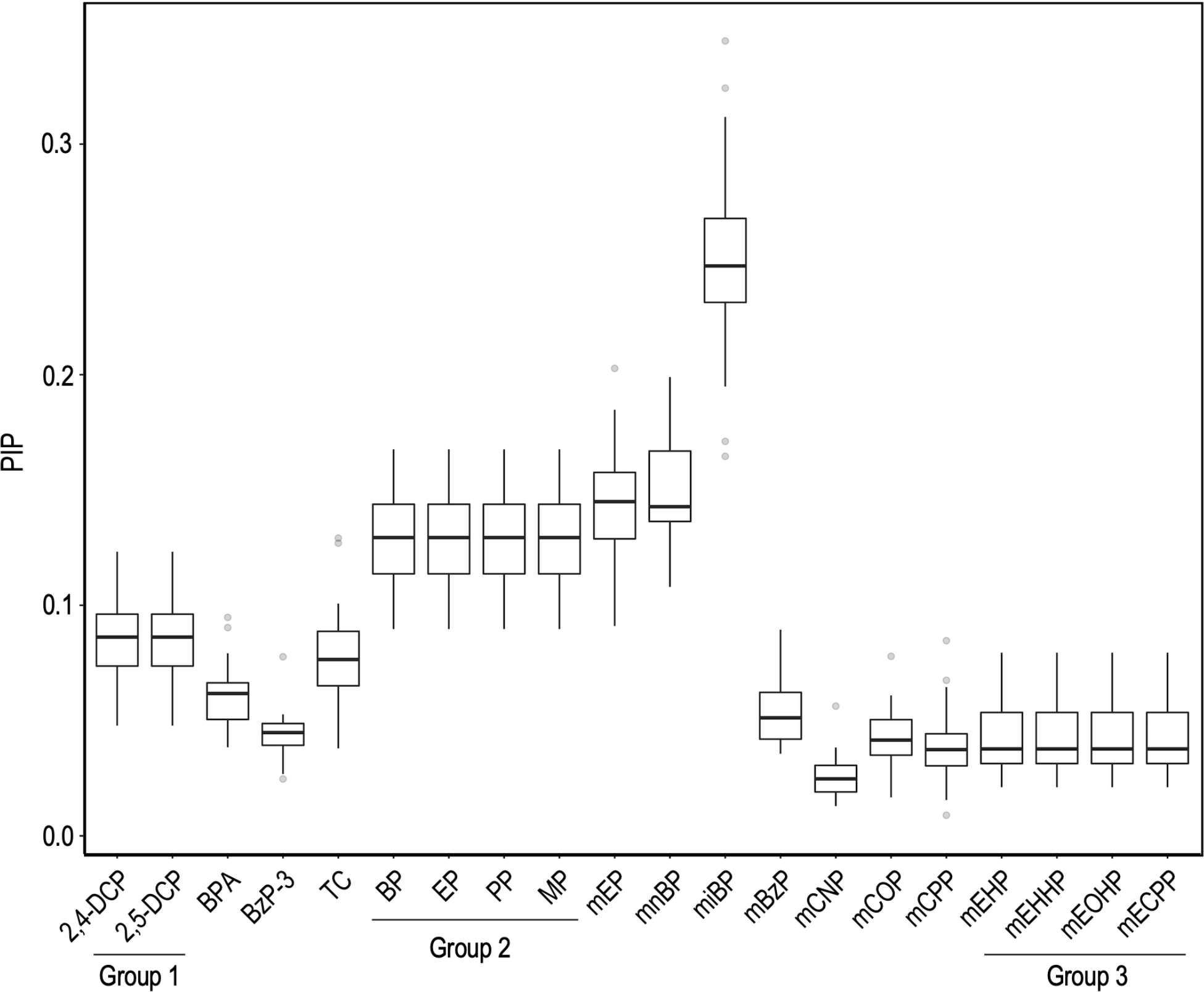
Individual posterior inclusion probabilities (PIP) for each ED metabolite or ED metabolites groups from BKMR. Abbreviations: Group 1 (2,4-dichlorophenol (2,4-DC), 2,5-dichlorophenol (2,5-DC)); bisphenol-A (BPA); benzophenone-3 (BzP-3); triclosan (TC); Group 2 (butyl-paraben (BP), ethyl-paraben (EP); propyl-paraben (PP); methyl-paraben (MP)); monoethyl phthalate (mEP); mono-n-butyl phthalate (mnBP); mono-isobutyl phthalate (miBP); monobenzyl phthalate (mBzP); monocarboxynonyl phthalate (mCNP); monocarboxyoctyl phthalate (mCOP), mono(3-carboxypropyl) phthalate (mCPP)), Group 3 (mono(2-ethylhexyl) phthalate (mEHP), mono(2-ethyl-5-hydroxy-hexyl) phthalate (mEHHP), mono(2-ethyl-5-oxohexyl) phthalate (mEOHP), mono(2-ethyl-5-carboxypentyl) phthalate (mECPP)).

**Fig. 4.**
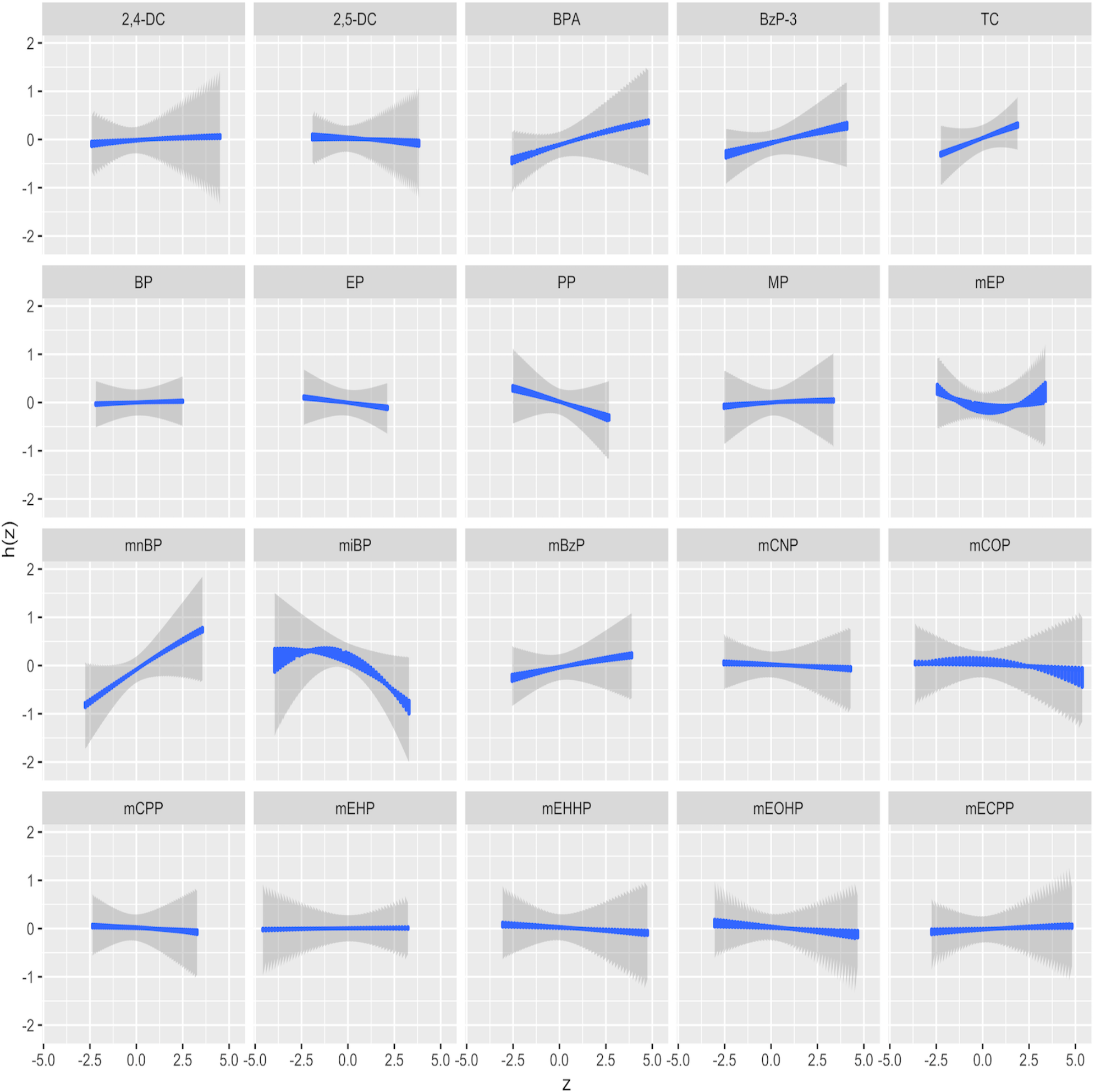
Univariate dose-response associations from BKMR. Plots assess the adjusted association between each ED exposure and the internalizing behaviour, while fixing the other ED exposures to their median. The estimate for each of the 30 multiply-imputed datasets are plotted and CI_95%_ are shown in grey to depict variability.

To better characterize the contribution of individual exposures to the overall effect, single-exposure effect estimates were computed (Fig. 5). Increases in individual exposure to 2,5-DC, Tc, parabens (BP, PP, MP), and a subset of phthalates (mnBP, miBP, mBzP) exhibited a trend for association with higher internalizing behaviour scores, when all of the remaining ED were fixed at their 75^th^ percentile as compared to when they were fixed at their 25^th^ percentile (Fig. 5). Again, CI for each compound estimate were wide across quantiles, indicative of an instability in the estimation. On the other hand, 2,4-DC, BPA, mCNP, and mCPP appeared not to contribute to the total effect on internalizing behaviour, as their mean TE estimates approached zero across the quantiles of concentrations (Fig. 5).

**Fig. 5.**
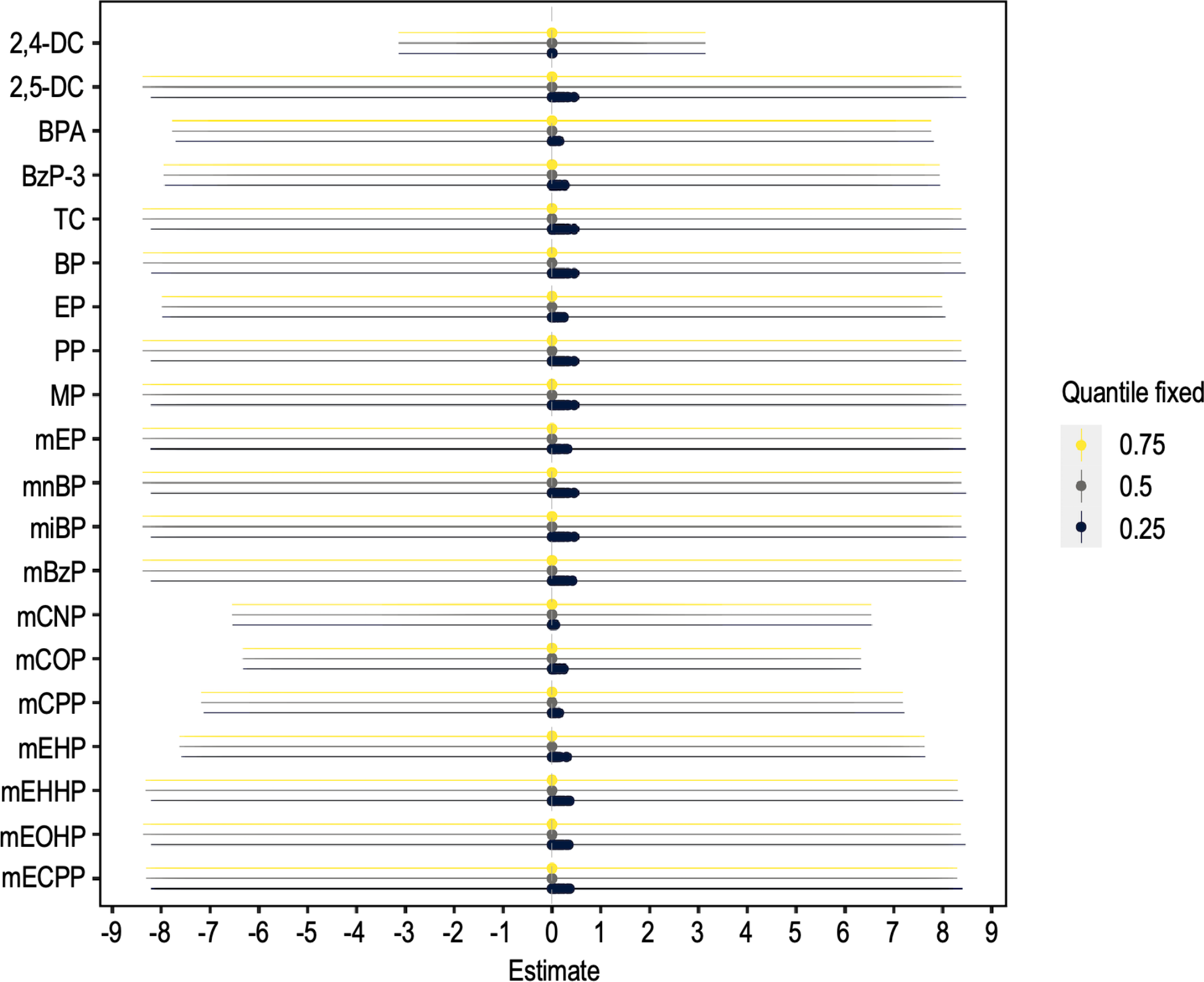
Single-exposure effects for each ED on internalizing behaviour score. Average change in internalizing score for a change in each of the 20 ED level from its 25^th^ to 75^th^ percentile, fixing the other ED levels at their 25^th^, 50^th^, or 75^th^ percentiles. The estimate for each of the 30 multiply-imputed datasets are plotted and the corresponding CI_95%_ are superimposed.

Despite the weak TE of ED mixture on internalizing scores and arguing that a suppressive effect of cytokines could have contributed to lower TE estimates, we decomposed the total effect of ED mixture exposure on internalizing behaviours into the NDE of ED mixture and the NIE mediated by each cytokine (Fig. 6, Tab. S1). For the different models, in the presence of either IL-1β, IL-6, or TNF-α, NDE effects were systematically lower than the TE (Fig. 6, Tab. S1). The NIE for each model including either IL-1β, IL-6, or TNF-α, were respectively 0.101, 0.1, and 0.102 (Fig. 6, Tab. S1), indicating that cytokines contribute to explain 10% of ED mixture TE on internalizing behaviours. However, CI_95%_ were very wide, again limiting the conclusions to be drawn. Our data therefore do not support a credible mediating or suppressive effect of cytokines of ED mixture exposure on internalizing behaviours.

**Fig. 6.**
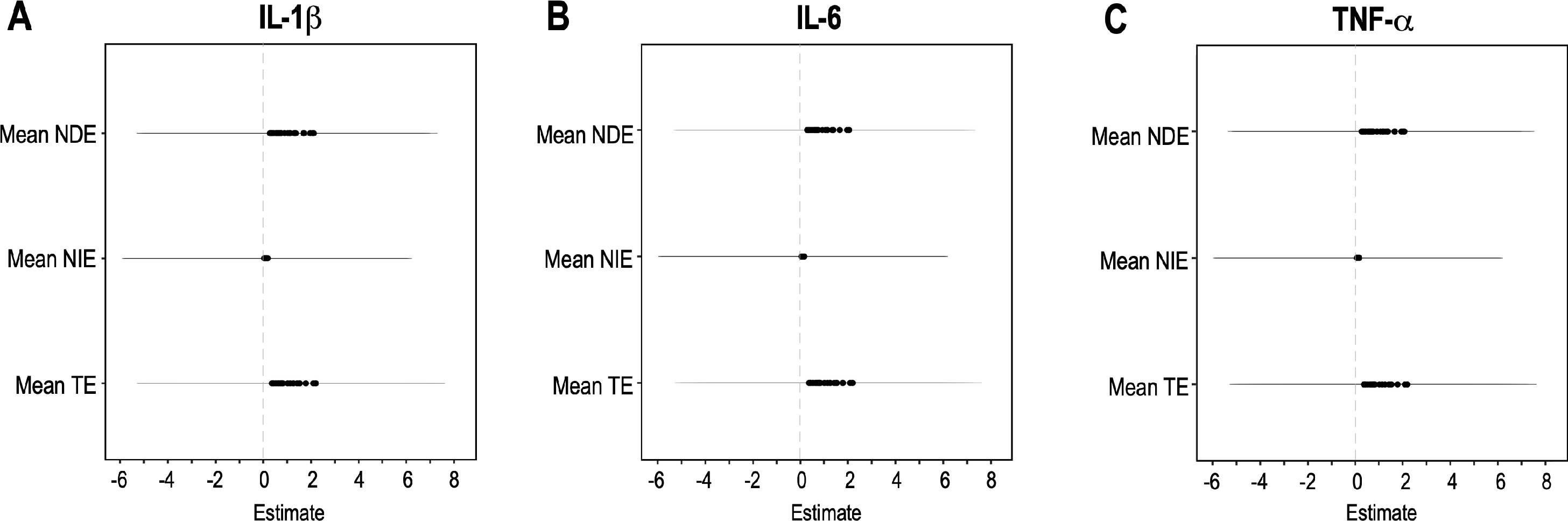
Natural direct effect of multiple ED exposure on internalizing behaviour and indirect effects of each cytokine. The total effect (TE) of multiple ED exposure was decomposed into the natural direct effect (NDE) and the natural indirect effect (NIE) for each cytokine of interest: IL-1β (A), IL-6 (B), and TNF-α (C). Individual dots represent the estimates for each of the 30 multiply-imputed datasets and the corresponding CI_95%_.

## 4. DISCUSSION

### Effects of prenatal exposure to ED mixture on internalizing behaviour at age 3 in boys

In this prospective birth cohort study and applying BKMR-CMA modelling, we found limited evidence to indicate that prenatal exposure to a complex mixture of 20 ED (5 phenols, 4 parabens, 11 phthalates and metabolites) negatively impacts neurodevelopment assessed by internalizing behaviours at age 3. Furthermore, the overall mixture effect was weak, with weak individual effect estimates and wide CI. The phthalates mnBP, miBP, and mEP appeared nevertheless to have the strongest influence, but their relatively modest PIP, as well as the wide CI of our estimates calls for caution in the interpretation of these results. Using a classical single pollutant modelling strategy, we previously found in the same study sample that individual exposure to mnBP was significantly associated with increased internalizing behaviour scores. Of note, none of the other 19 ED considered was significantly associated with increased internalizing behaviour score. Moreover, mnBP effects were weak (OR=1.06; CI_95%_ [1.01;1.11]) and did not survive multiple testing (Philippat et al., 2017). This argues in favour of a generally weak effect of this ED mixture on internalizing behaviour in this study sample, possibly driven by the phthalate mnBP.

It remains difficult to compare our results to other studies, as scarce studies have addressed ED mixture exposure effects on internalizing behaviours in the general population. Also, non-overlapping lists of ED are generally considered in mixture. For example, we show that increased exposure to an ED mixture (consisting of 12 phenols, 13 phthalates, 2 non-phthalate plasticizer metabolites) was associated with higher internalizing behaviour scores, applying Weighted Quantile Sum (WQS) regression io a study sample of 416 mother-child pairs from the French SEPAGES cohort (Guilbert et al., 2021). The highest weights were observed for mEP, mBzP, and mnBP. This is partially in agreement with our BKMR analysis in the EDEN cohort as PIP, univariate dose-response associations and single-exposure effect pointed towards mnBP as a possible driver of ED mixture effect on internalizing behaviour score. However, the ED mixture considered by Guilbert was different from our study and ED levels were measured in within-subject pools of multiple urine samples collected at two distinct time points during pregnancy (>20 samples/individual pool), resulting in a more accurate exposure measure. Still, the score increase in girls was 2.47 points (CI_95%_ [0.6;4.33]) for an increase of one tertile in the WQS index, compared with 1.17 points (CI_95%_ [-0.5;2.84]) in boys. Considering the wide CI encompassing zero in boys, this study suggests a weak or absence of effects of ED mixture exposure in boys. Therefore, focusing only in boys and relying only on a single ED mixture exposure measure point may have contributed to weaken the effects we observe in the present study.

We cannot exclude the fact that our results may actually reflect a lack of effects of ED mixture exposure on internalizing behaviours. This would be in agreement with a previous report of a lack of association between prenatal exposure to a mixture of phthalates, bisphenol, and organophophorous pesticides with internalizing or autistic behaviours at age 6 in 782 children from the Generation R cohort (van den Dries et al., 2021). Also, no association between prenatal mixture exposure to the phthalates mnBP, mBzP, miBP, mEP, and four DEHP metabolites and internalizing behaviours or anxious-shy behaviours was observed among 411 mother-child pairs (Daniel et al., 2020). Quite interestingly, an associations with anxious-shy behaviours was observed only when considering mixture exposure to a specific subset of DEHP metabolites (Daniel et al., 2020). As stressed by the authors, the latter results advocate that the inclusion of compounds with distinct biological activity in the same models (for example DEHP and non-DEHP metabolites) may bias towards the null hypothesis and may call for favouring mixtures of ED with common biological activities. In our case, we included 20 ED from three main families of compounds known to act on distinct biological pathways. This may have contributed to hinder the true biological effect of specific ED present in our mixture on internalizing behaviours.

### ED mixture exposure and other neurodevelopmental outcomes

When considering other outcomes related to neurodevelopment, mixed reports coexist in the literature. One recent study reported that prenatal exposure to an ED mixture of BPA, Tc, phthalates, and perfluorinated compounds (partly overlapping with our 20 ED mixture), was associated with higher odds of language delay in a sample of more than 2000 infants (Caporale et al., 2022). Also, higher prenatal exposure to a mixture of phthalates, bisphenols, and organophosphorous pesticides was associated with lower nonverbal intellectual quotient at age 6 in 782 children from the Generation R cohort (van den Dries et al., 2021). Another study found evidence that prenatal exposure to a mixture of organophosphorous pesticides increased the odds of ASD in a cohort of 237 typically developing and 224 children with ASD (Bennett et al., 2022). On the other hand, another study provided limited evidence for adverse relationship between prenatal exposure to a mixture of dioxins and polychlorinated biphenyls with psychomotor development or mental development at 6 months of age in a sample of 259 infants (Yim et al., 2022). These studies suggest that depending on the ED mixture (complexity, diversity, measure mode), the psychopathological outcome, or the population sample (clinical, general population, boys only), distinct conclusions can be drawn related to the adverse effects of ED mixture exposure on child’s behaviour in general.

### Mediation or suppression of ED mixture effects on internalizing behaviour by cytokines

Since an association between a specific ED mixture and health outcome does not necessarily mean that the relationship is causative, identifying possible intermediary pathways underlying ED mixture effects would be one move towards causality or at least towards an understanding of the underlying biological pathways. The recent development of BKMR-CMA provides methodological framework to address this. To our knowledge, no other study has addressed whether perinatal inflammation could be a plausible biological pathway which could modulate the effects of ED mixture exposure on neurodevelopment. Although our results indicated that the total effect of ED mixture exposure on internalizing behaviours was at best weak or absent, we considered the mediation analysis could uncover a suppressive effect of cytokines, which could have contributed to lower the total effect estimates. This was not the case and given the wide credible intervals for the mediation estimates, our analyses suggest that cytokines elevated in inflammatory conditions (IL-1β, IL-6, TNF-α) are unlikely mediators of the deleterious effects of ED mixture exposure on internalizing behaviours.

### Limitations of the study

Although our study sample was well characterized in terms of covariates which could influence either the exposure, outcome or mediators, this study also presents some limitations. First, although the window of sampling was narrow (25-29 weeks amenorrhea), ED were measured at a single time point in maternal urines, this might not reflect accurately long-term ED foetal exposure as placental barrier permeability to the different classes of ED may vary. Second, as statistical strategies to model ED mixture exposures effects complexify, improving power by relying on a larger population sample may be critical to grasp complex effects of ED mixture. Third, we have only addressed the effects of ED exposure mixture on internalizing behaviours and have not considered neuropsychopathological outcomes more proximal to the exposure window. Fourth, we have only considered boys and our results have not been replicated yet in an independent cohort, precluding generalization of our findings.

### How to improve future studies?

Recent development of powerful analytical strategies now allows estimation of ED mixture effects on human health in general, and on neurodevelopment in particular. It may be timely to set high standards for such analyses to improve reliability of this emerging literature and to leave room for reports of weak or lack of effects. In terms of standards, relying on population samples of larger sample size, contrasting results obtained from boys and girls, improving the accuracy of ED exposure measures by multiple sampling appear critical to capture the adverse effects of ED mixture on neurodevelopment. Also, to guide accurate hypothesis formulation as well as the ED mixture composition, it is important to account for prior reports of weak or low association and *a priori* knowledge related to each ED biological activity. Indeed, it is plausible that considering small sample size and ED mixtures compounds with distinct biological activity in the same models may bias towards the null hypothesis. These considerations are in line with a recent review (Joubert et al., 2022), which advocates for the use of the most recent statistical advancements for data analysis to improve exposure-response estimation, for increased accuracy of exposure measures, this to globally enable more informed analyses of environmental mixtures effects. Finally, identifying biological pathways through mediation studies should highlight targets for prevention. Pursuing in this direction should improve risk assessment of ED mixture exposures on child’s neurodevelopment to reduce exposure and improve public health.

## 5. CONCLUSIONS

Our study provides limited evidence supporting a deleterious impact of prenatal exposure to a mixture of phenols, parabens, and phthalates on internalizing behaviours in boys at age 3. Also, the pro-inflammatory cytokines IL-1β, IL-6, and TNF-α are unlikely mediators of the detrimental effect of prenatal exposure to ED mixture. Although this could be interpreted as a weak or null effect of this ED mixture exposure and cytokines on neurodevelopment, it is possible that our study design (sample size, single exposure and mediator assessment, considering boys only) may have contributed to hinder the actual true effect of ED mixture exposure on internalizing behaviour. As mixture and mediation methods are continuously evolving, future studies should set best practices both in terms of biostatistical strategy but most importantly in terms of study design to reliably estimate the actual true effect of prenatal ED mixture exposure on neurodevelopment and guide public health policies to regulate ED mixture exposures.

## Data Availability

Our dataset will be made available by the corresponding author upon reasonable request and acceptance by the EDEN birth cohort steering committee.

## List of abbreviations

2,4-DC: 2,4-dichlorophenol
2,5-DC: 2,5-dichlorophenol
ASD: autism spectrum disorder
BKMR-CMA: Bayesian Kernel Machine Regression Causal Mediation Analysis
BP: butyl-paraben
BPA: bisphenol A
BzP-3: benzophenone-3
DEHP: di(2-ethylhexyl) phthalate
ED: endocrine disruptor
EP: ethyl-paraben
IL: interleukin
mBzP: monobenzyl phthalate
mCNP: monocarboxynonyl phthalate
mCOP: monocarboxyoctyl phthalate
mCPP: mono(3-carboxypropyl) phthalate
mECPP: mono(2-ethyl-5-carboxypentyl)
mEHHP: mono(2-ethyl-5-hydroxy-hexyl) phthalate
mEHP: mono(2-ethylhexyl) phthalate
mEOHP: mono(2-ethyl-5-oxohexyl) phthalate
mEP: monoethyl phthalate
miBP: mono-isobutyl phthalate
mnBP: mono-n-butyl phthalate
MP: methyl-paraben
PP: propyl-paraben
Tc: triclosan
TNF: Tumor Necrosis Factor
PIP: Posterior Inclusion Probability

## FUNDING SOURCES

NG and RS are grateful to the Agence nationale de sécurité sanitaire de l’alimentation, de l’environnement et du travail ANSES (“Environnement-santé-travail” funding scheme) for funding the present research. LD, CG, and BH thank the the Fondation de France (“Autism and neurodevelopment” funding scheme) for funding the present research. The funders played no role in the design and conduct of the study; the collection, management, analysis, and interpretation of the data; the preparation, review, and approval of the manuscript; or the decision to submit the manuscript for publication.

## ETHICS APPROVAL

The EDEN cohort promoted by INSERM has received approval from the CCPPRB (Comité consultatif de protection des personnes dans la recherche biomédicale, the French Institutional Ethic Board for human subjects biomedical research) of Kremlin Bicêtre and from the CNIL (Commission Nationale Informatique et Liberté, the French national agency on data privacy) for biological samplings and data collection. Informed written consent was obtained from parents at the time of enrolment and after delivery. The use of EDEN samples and data in the present study has been approved by the EDEN steering committee. The analysis of blinded urine specimens at CDC laboratories was determined not to constitute engagement in human subjects’ research.

## Appendix A. SUPPLEMENTARY INFORMATION TO

### SUPPLEMENTARY FIGURES

**Fig. S1.**
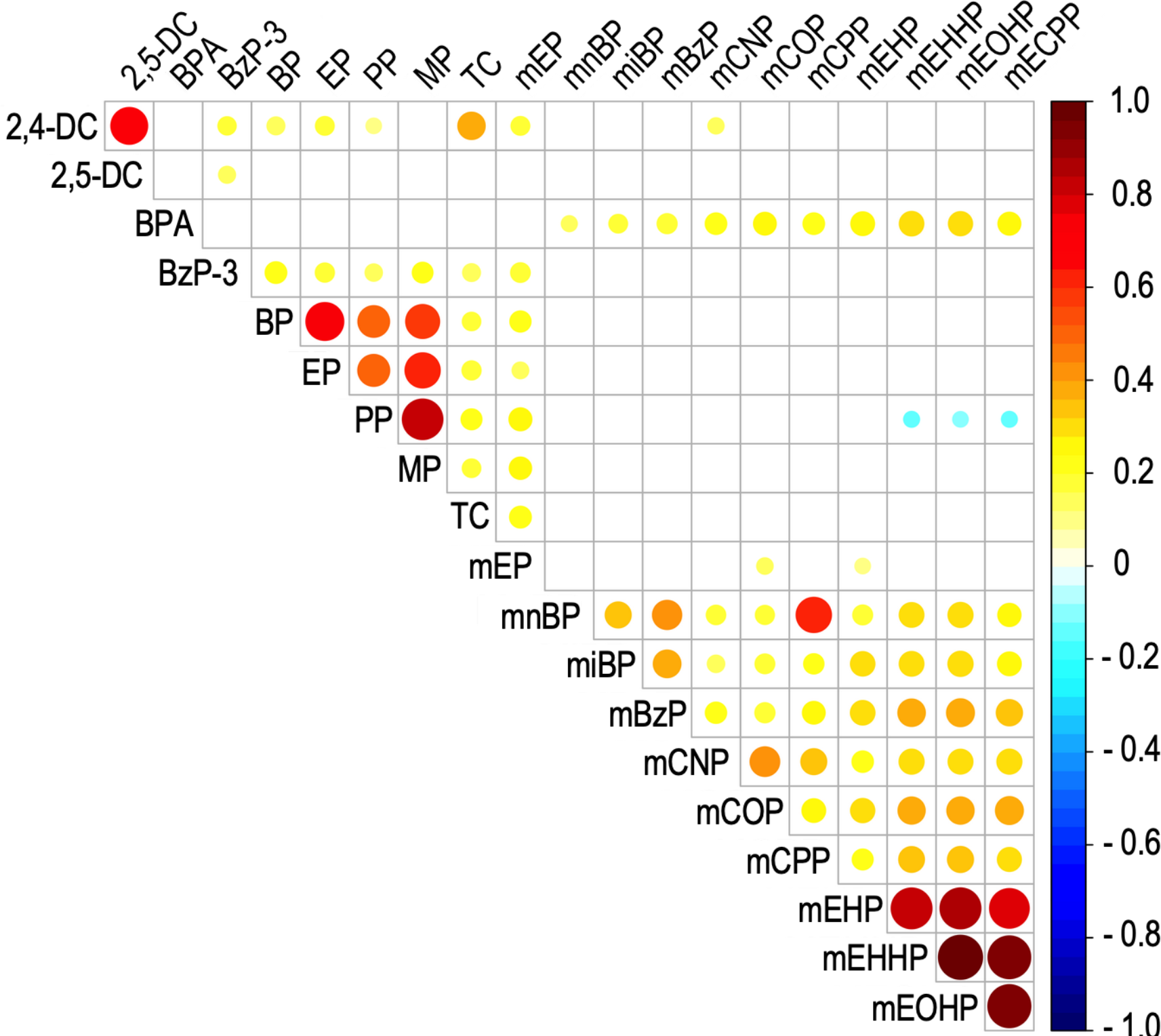
Correlation analysis of ED levels in maternal urinary samples. Heatmap of the pairwise Spearman’s rank rho correlation coefficient between all ED pairs. Spearman’s rho coefficients are color-coded and proportional to dot area. Only significant correlations are displayed (adjusted p-value < 0.05).

**Fig. S2.**
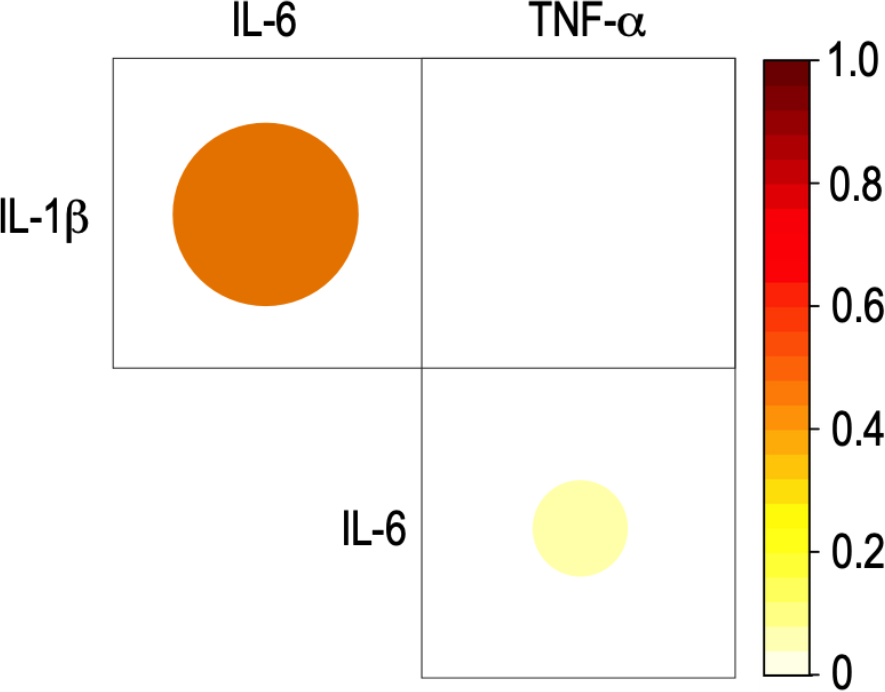
Correlation analysis of the serum levels of IL-1β, IL-6, and TNF-α in cord blood samples. Heatmap of the pairwise Spearman’s rank rho correlation coefficient between all cytokine pairs. Spearman’s rho coefficients are color-coded and proportional to dot area. Only significant correlations are displayed (adjusted p-value < 0.05).

### SUPPLEMENTARY TABLE

**Tab. S1.** Mediation analysis in the BKMR framework using the 20 ED mixture as exposure and each cytokine as the mediator. Pooling of the estimates over 30 MI datasets was performed using the mean and SD. TE, total effect; NDE, natural direct effect; NIE, natural indirect effect.

| | IL-1β | | IL-6 | | IL-10 | | TNF-α | |
| --- | --- | --- | --- | --- | --- | --- | --- | --- |
| Estimates | Mean | SD | Mean | SD | Mean | SD | Mean | SD |
| TE | 1.018 | 2.840 | 1.018 | 2.840 | 1.018 | 2.840 | 1.018 | 2.840 |
| NDE | 0.916 | 2.750 | 0.920 | 2.770 | 0.917 | 2.770 | 0.920 | 2.790 |
| NIE | 0.101 | 2.920 | 0.100 | 2.940 | 0.100 | 2.940 | 0.102 | 2.960 |

## Notes

### Competing Interest Statement

The authors have declared no competing interest.

### Funding Statement

NG and RS are grateful to the Agence nationale de sécurité sanitaire de l'alimentation, de l'environnement et du travail ANSES (Environnement-santé-travail funding scheme) for funding the present research. LD, CG, and BH thank the the Fondation de France (Autism and neurodevelopment funding scheme) for funding the present research. The funders played no role in the design and conduct of the study; the collection, management, analysis, and interpretation of the data; the preparation, review, and approval of the manuscript; or the decision to submit the manuscript for publication.

### Author Declarations

The EDEN cohort promoted by INSERM has received approval from the CCPPRB (Comité consultatif de protection des personnes dans la recherche biomédicale, the French Institutional Ethic Board for human subjects biomedical research) of Kremlin Bicêtre and from the CNIL (Commission Nationale Informatique et Liberté, the French national agency on data privacy) for biological samplings and data collection. The use of EDEN samples and data in the present study has been approved by the EDEN steering committee.

